# Blood folate level needed for fully effective neural tube defect prevention

**DOI:** 10.1101/2024.09.11.24313482

**Authors:** Nicholas J. Wald, Stephen H. Vale, Jonathan P. Bestwick, Joan K. Morris

## Abstract

**Introduction:** Neural tube defects (NTDs) are a folate deficiency disorder. NTDs are preventable; increasing folic acid intake through food fortification or use of supplements increases serum and red blood cell folate and reduces the risk of a woman having an NTD pregnancy. There is controversy over whether there is a blood folate level needed to achieve the full preventive effect because of discrepant conclusions from studies investigating the relationship between folate levels and NTD risk. Resolving the controversy is important in determining public health policy.

**Methods:** Data from two studies conducted in Ireland and China were used to determine the relationship between serum folate and NTD risk. The relationship from each study was compared with the observed result in a randomised trial of folic acid that increased serum folate from 5 ng/ml to 44 ng/ml among the women who took the daily folic acid supplement before and during early pregnancy.

**Results:** Data from both studies showed a proportional (logarithmic) relationship between serum folate and NTD risk with no evidence of a folate threshold above which there is no further NTD risk reduction. Both studies accurately predicted the observed result from the randomised trial that achieved serum folate levels beyond the average levels found in the general population with an 83% preventive effect. This is higher than can be achieved with current levels of folic acid food fortification or with the recommended peri-conceptional use of 0.4mg supplements. The suggestion of a threshold is not due to discrepant data but the incorrect interpretation of the folate-NTD risk association when plotted on arithmetic scales which conceals the proportional relationship between the two.

**Conclusion:** To achieve fully effective fortification serum folate levels need to be about 44ng/ml.

**Key Messages:** *What is already known on this topic:* Neural tube defects (NTDs), among the most common serious birth defects worldwide, are a folate deficiency disorder. Increasing folic acid (vitamin B9) intake increases serum folate and reduces the risk of an NTD pregnancy. There is a difference of opinion on the serum folate level needed for fully effective NTD prevention.

*What this study adds:* There is no threshold above which NTD risk does not decrease and population folate levels need to be substantially increased to have the expected potential achieve effect on the prevention of NTDs.

*How this study might affect research, practice or policy:* A serum folate level of about 44ng/ml is a reasonable target to achieve an approximate 83% reduction in the prevalence of NTD pregnancies. This result can be used to help influence folic acid fortification policy.

## INTRODUCTION

Neural tube defects (NTDs) are one of the most serious and common birth defects throughout the world. It is recognized that NTDs are a folate deficiency disorder. Increasing folic acid intake through food fortification or the use of supplements increases serum and red blood cell (RBC) folate and reduces the risk of a woman having an NTD pregnancy. There is, however, a lack of clarity over the blood folate level needed to achieve the full preventive effect. Two estimates have been published, one by Wald et al.^**1**^ (2001) based mainly on data from Daly et al.^**2**^ (1995) and the other by Crider et al.^**3**^(2014) that appear discrepant, with Daly et al.^**2**^ and Crider et al.^**3**^ suggesting a blood folate threshold above which there is no further NTD risk reduction and Wald et al. ^**1,4**^ suggesting there is no threshold.

In Daly et al.^**2**^ results were from a large observational cohort study that quantified the relationship between serum and RBC folate and NTD risk. In 1998 it was shown that the relationship described by Daly et al.^**2**^ between RBC folate and NTD risk was linear when RBC folate and NTD risk were both expressed in logarithms^**4**^ and the same is true of serum folate showing that a doubling of serum folate approximately halves the risk of having an NTD pregnancy^**1**^. For example, increasing serum folate levels from a typical background level of 5ng/ml to 10ng/ml would be expected to halve NTD risk. The analysis showed that the effect of increasing serum folate on predicted NTD risk depended on baseline folate levels and the slope of the log-log serum folate-NTD risk relationship with no serum folate threshold above which there is no further risk reduction.

In 2014 Crider and colleagues^**3**^ using data from two studies carried out in China estimated the relationship between red cell folate and NTD risk and concluded that there was a threshold at an RBC folate level of about 1000nmol/l (441ng/ml). Daly et al.^**2**^ had reached a similar conclusion, with a threshold at an RBC folate level of about 1300 nmol/l (574 ng/ml).

We here explore the basis for the long standing difference in opinion on the issue of whether or not there is a threshold and the implications on folic acid fortification policy.

## MATERIALS AND METHODS

The relationship between log serum folate levels and log NTD risk were plotted using the Daly et al. ^**2**^ results allowing for regression dilution bias as was done in Wald et al. ^**1**^ The Crider et al.^**3**^ results were based on RBC folate levels that were converted into serum folate results using data from Chen et al.^**5**^ Both of these ‘dose-response’ relationships were tested to see how well they predicted the observed result among non-pregnant women who took 4mg folic acid daily in a randomised trial published in 1991 (Wald et al. MRC Vitamin Study^**6**^) which resulted in a risk reduction of 83% (on-treatment analysis) as a result of an observed increase in the median serum folate from 5ng/ml at baseline to 44ng/ml, an 8.8-fold increase. The detailed calculations are given in the statistical appendix.

## RESULTS

Figure 1 shows the relationship between serum folate and NTD risk, both plotted on log-scales based on the results of Daly et al. ^**2**^ and Wald et al.^**1**^ (labelled Daly/Wald). The solid “dose-response” line represents the range over which there were observed data from Daly et al.^**2**^ The extrapolation of this line (dotted) shows how well the dose-response relationship predicts the observed results in the report of the MRC Vitamin Study trial^**6**^ well beyond the range of serum folate levels observed by Daly et al. ^**2**^ The predicted risk reduction for an increase in serum folate from 5 to 44 ng/ml is 83% (See Section 1 of the statistical appendix). The prediction is identical to the observed risk reduction in the MRC Vitamin Study^**6**^.

**Figure 1.**
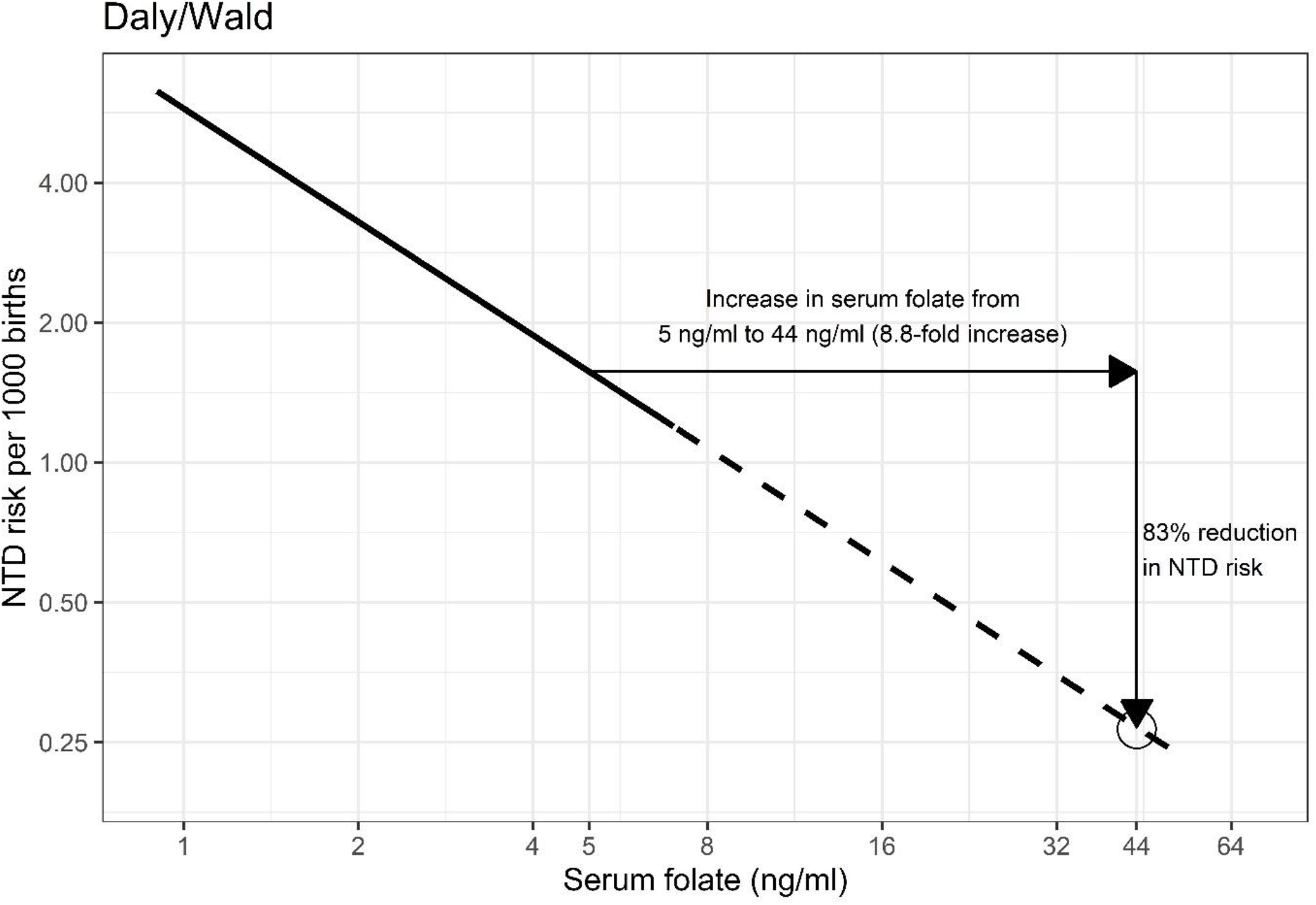
Relationship between serum folate and neural tube defect (NTD) risk based on data from Daly et al. ^**2**^ allowing for regression dilution bias. The circle indicates the reduction in NTD risk observed in the MRC Vitamin Study (1991) ^**6**^. The full line corresponds to the range of results given in Daly et al. ^**2**^ and the dotted line extrapolates the results to higher serum folate levels.

Figure 2, derived from Chen et al. ^**5**^, is a plot of log RBC folate against log serum folate. There is a log-log linear relationship which was used to convert Crider et al’s ^**3**^ RBC results into equivalent serum folate results and used in Figure 3. Figure 3 (labelled Crider/Chen) shows the relationship between serum folate derived from RBC folate and NTD risk both plotted on log-scales. Since the relationship is modelled the regression line is dotted. The predicted risk reduction is 82% for an increase from 5 to 44ng/ml in serum folate (8.8-fold increase, see Section 2 of the statistical appendix) again a nearly perfect prediction of the observed risk reduction in the MRC Vitamin Study^**6**^.

**Figure 2:**
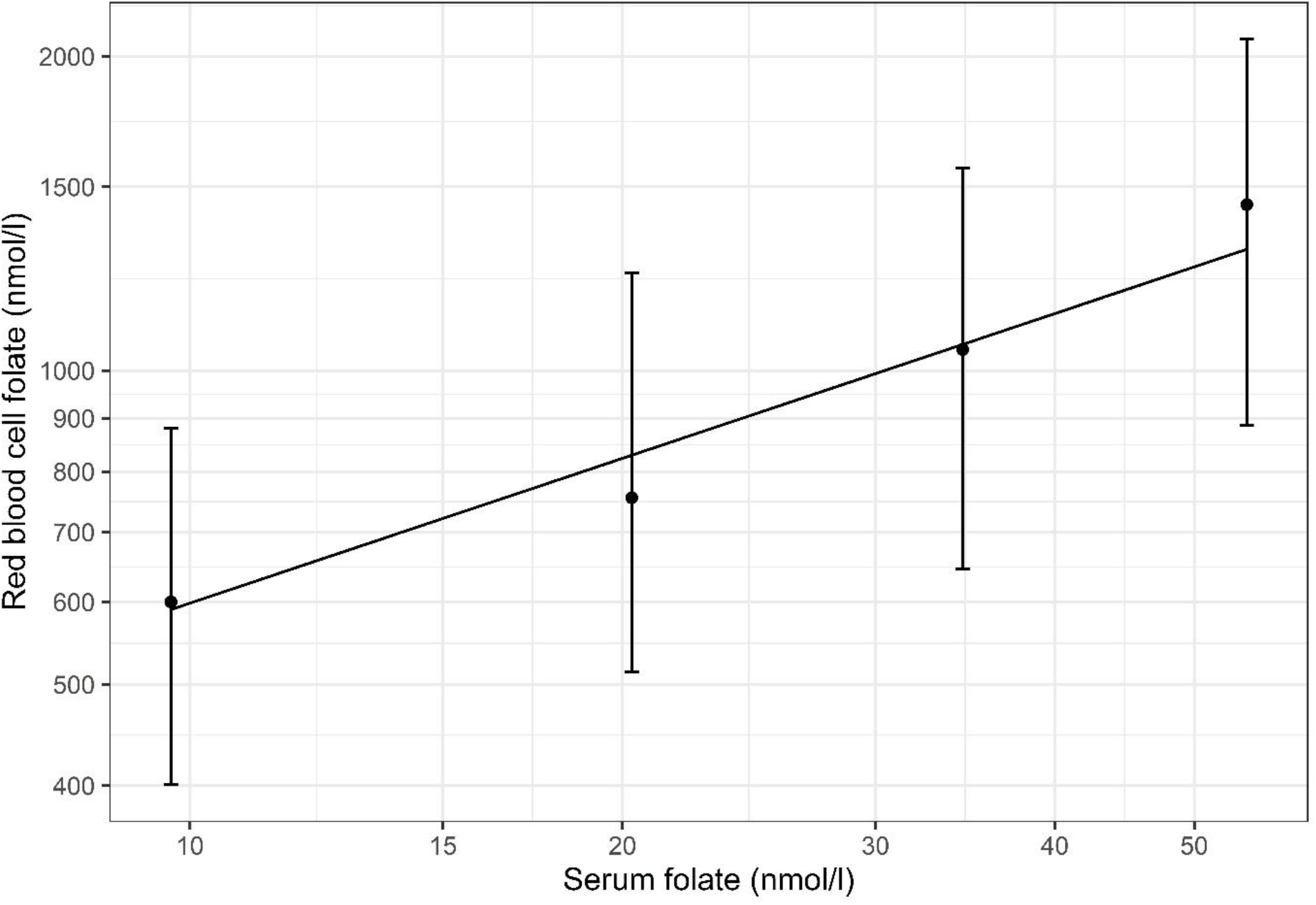
log-log linear regression between serum folate levels and red blood cell (RBC) folate levels derived from data in Supplemental Table 1 in Chen et al.^**5**^. The RBC folate level was regressed against serum folate level for the baseline and folic acid doses of 100, 400 and 4000 μg per day given in Supplemental Table 1 of Chen et al. ^**5**^

**Figure 3:**
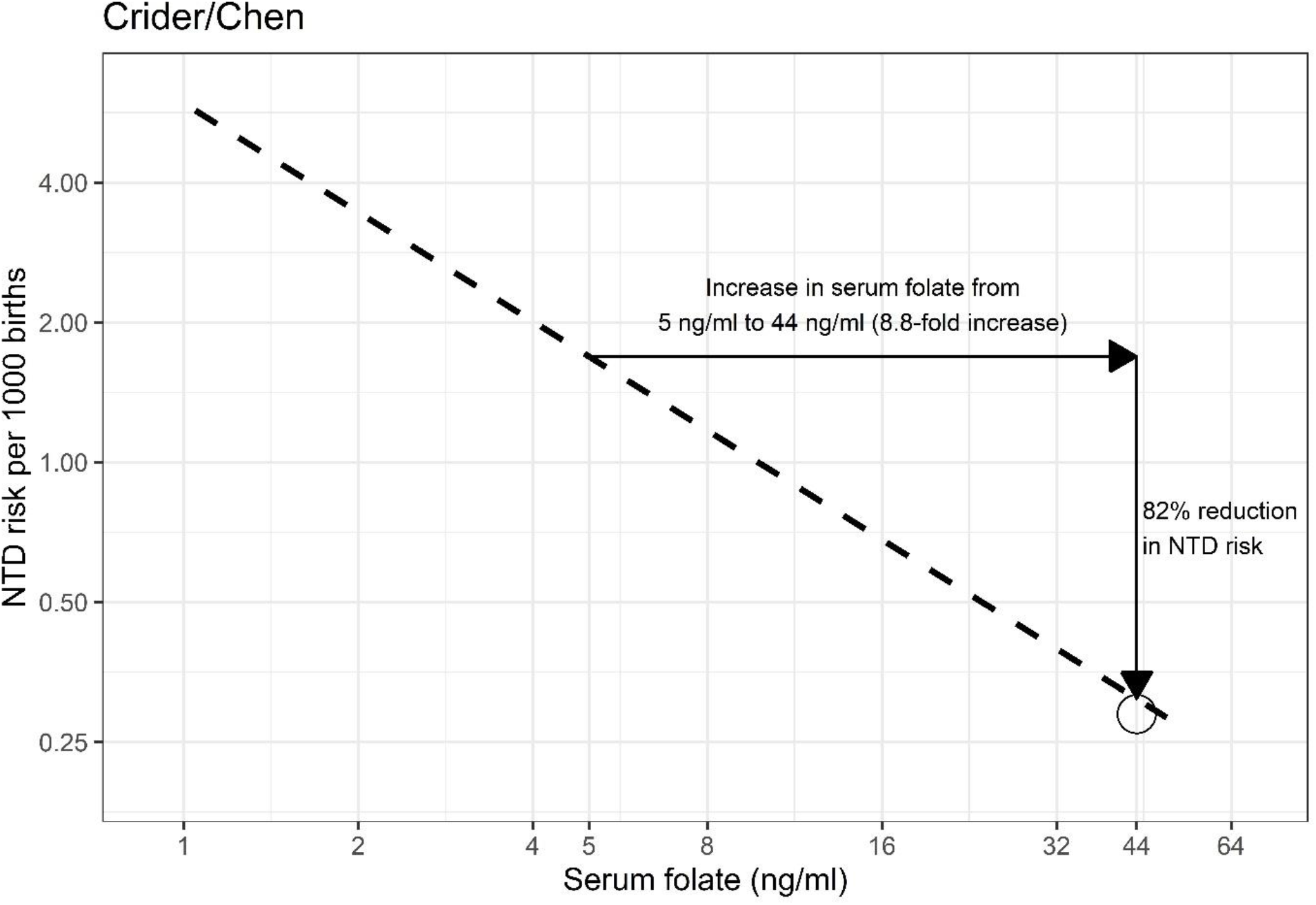
Relationship between serum folate (derived from RBC folate) and NTD risk (Crider et al. ^**3**^). The circle indicates the reduction in NTD risk observed in the MRC Vitamin Study (1991) ^**6**^.

Figures 4 (a) and (b) display Figures 1 and 3 respectively on arithmetic scales. They show how failure to use proportional (i.e. log) scales creates the false impression that the effect of blood folate on NTD risk tapers off to nil, implying a threshold.

**Figure 4:**
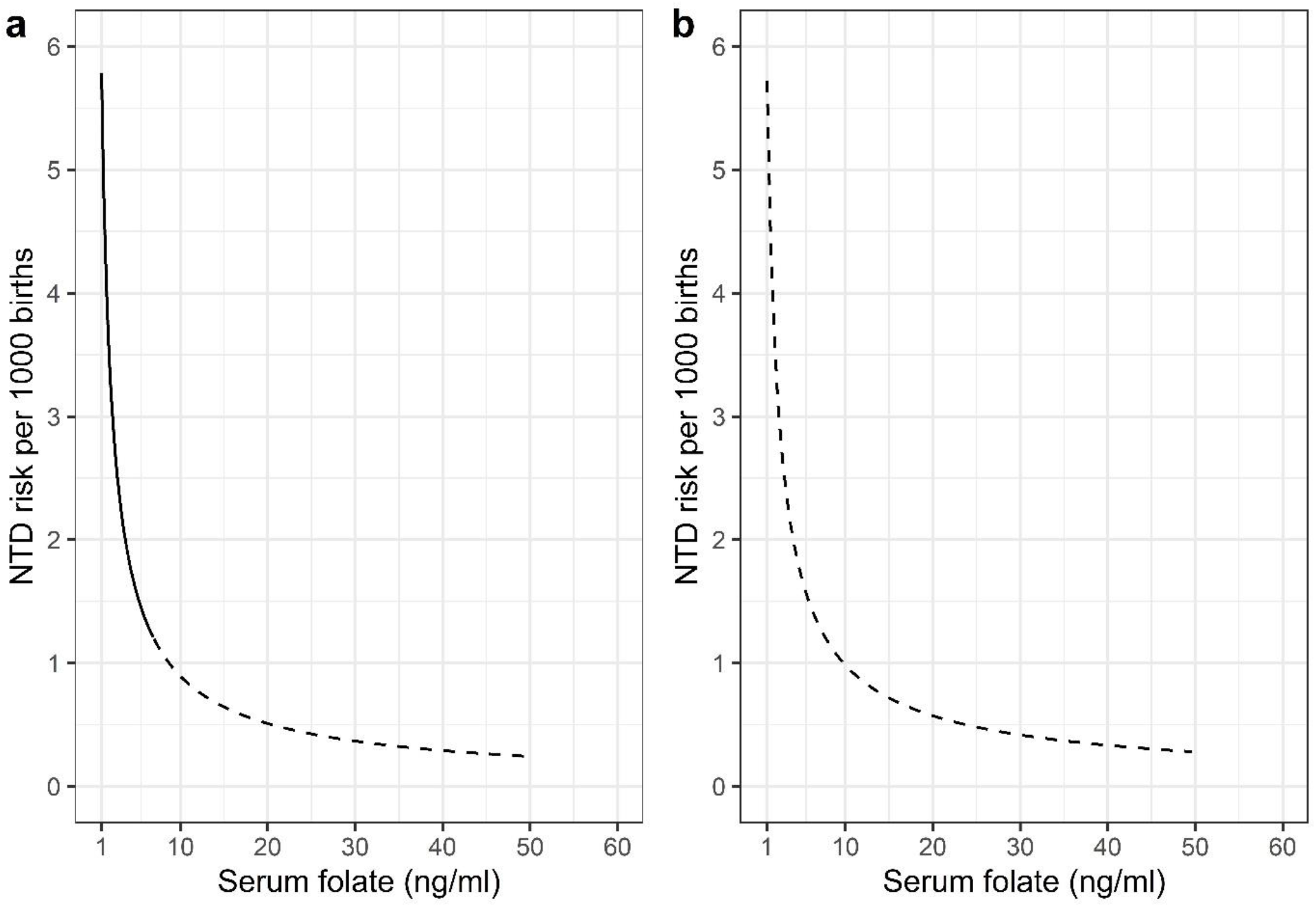
Relationship between serum folate and NTD risk plotted on arithmetic scales for Daly et al. ^**2**^ (Figure 4a) and for Crider et al.^**3**^ (Figure 4b) using RBC folate measurements converted to serum folate measurements. The full line corresponds to the range of results with observed data (Daly et al. ^**2**^)

## DISCUSSION

Three important conclusions can be drawn from our results. First, there is no discrepancy between the predicted risk reductions based on the two data sets, Daly/Wald and Crider/Chen; the two dose-response slopes are virtually identical. Second, neither dose-response relationship shows a threshold in which an increase in blood folate level ceases to reduce NTD risk. Third, the false perception that there is a threshold stems from displaying the results of the relationships on arithmetic scales even though the statistical analyses were based on logarithms. This false perception has had significant public health implications.

Crider et al.^**3**^ show good agreement between their data and that of Daly et al.^**2**^ but this is limited to the narrow range of serum folate levels observed in Daly et al.^**2**^ Our paper shows that the agreement persists at serum folate levels well beyond those observed in Daly et al.^**2**^ and that both studies predict the observed risk reduction in the MRC Vitamin Study^**6**^.

The proportional relationship between serum folate and NTD risk may give the impression of a threshold when plotted on arithmetic scales (Figure 4) because the same proportional reduction will result in smaller absolute reductions as the NTD prevalence decreases. As the log-log plots show there is no indication of a threshold above which an increase in blood folate level ceases to reduce NTD risk and consequently there is no evidence of two mutually exclusive categories of potential NTD pregnancies, one folic acid responsive and another which is not folic acid responsive.

Countries that have mandated folic acid fortification policy have done so insufficiently, resulting in increases in serum folate too small^**7,8**^ to achieve fully effective fortification. Until fully effective fortification is implemented all women who may become pregnant should be advised to take a daily folic acid supplement of 4mg (or 5mg if 4mg is not available) instead of 0.4mg, but folic acid fortification designed to achieve a median serum folate level of about 44 ng/ml is the policy of choice. It obviates the need to take folic acid supplements which is, anyway, of limited effectiveness because most women do not take supplements when it is needed, immediately before pregnancy^**9**^.

As well as estimating the serum folate level needed for fully effective fortification attention needs to be paid to the issue of safety. There is no evidence or indication that serum folate levels of 44 ng/ml pose any risk to health whereas not achieving fully effective fortification will knowingly cause harm^**10**^.

The term “fully effective fortification”, strictly interpreted, implies a serum folate threshold. The term can also be used to set a target serum folate level that was directly observed in a randomised clinical trial that resulted in a large preventive effect, above which there is minimal extra prevention. We used the term in this sense. This alone supports the conclusion that 44 ng/ml be considered a target level for serum folate.

The World Health Organization has estimated that, globally every year, there are more than 300,000 NTD pregnancies^**11**^. The fortification policy targeted at a median serum folate level of 44 ng/ml could prevent about a quarter of a million cases every year (83% of 300,000). It would achieve a major public health benefit to otherwise affected individuals, their families and society.

Public health authorities should consider implementing fully effective fortification with serum folate monitoring among samples of the population to ensure the fortification policy is achieving its full potential in preventing one of the most serious and common birth defects throughout the world.

## Data Availability

All data produced in the present work are contained in the manuscript

## STATISTICAL APPENDIX

All logarithms are to the base *e*.

### 1. Daly/Wald

i. The results of Daly et al.^**2**^ and Wald et al.^**1**^ show that there is a log-log relationship between NTD risk and serum folate level with slope −0.81 when corrected for regression dilution bias. The relationship is:

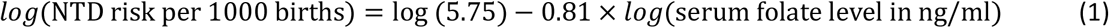

The value of 5.75 NTDs per 1000 births was taken from Daly et al.^**2**^ from their observation that the median serum folate level corresponded to 1.9 NTDs per 1000 births.
ii. For a log-log relationship between NTD risk and serum folate level:

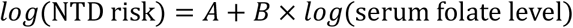

(serum folate level) If the serum folate level increases from *SF*_1_ to *SF*_2_the percentage NTD risk reduction will be:

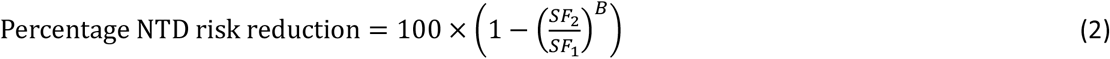
iii. The predicted decrease in risk using the results from Daly/Wald can be found by substituting an 8.8-fold increase in serum folate level and the value of *B* in Equation (1) into Equation (2):

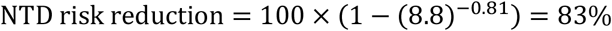

### 2. Crider/Chen

i. The results in Supplemental Table 1 of Chen et al.^**5**^ were used to show that there is a log-log linear relationship between serum folate level and red blood cell (RBC) folate level. A log-log regression of the RBC folate level against serum folate level for the baseline and folic acid doses of 100, 400 and 4000 μg per day in Supplemental Table 1 gives the following relationship:

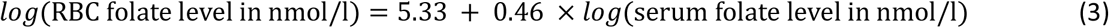
ii. The results in Supplemental Table F of Crider et al^**3**^ show that there is a log-log relationship between NTD odds and serum folate level. Assuming NTD odds and NTD risk are approximately equal because the NTD risk is small:

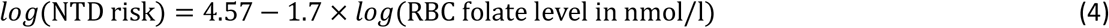

Substituting Equation (3) in Equation (4) gives the following relationship between NTD risk and serum folate level:

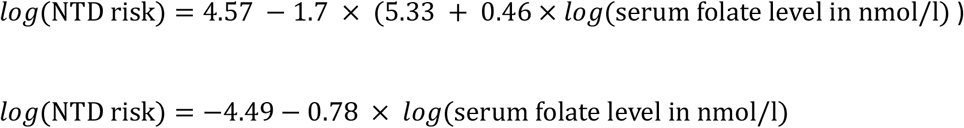

For consistency with Equation (1), convert NTD risk to NTD risk per 1000 births and serum folate level to ng/ml:

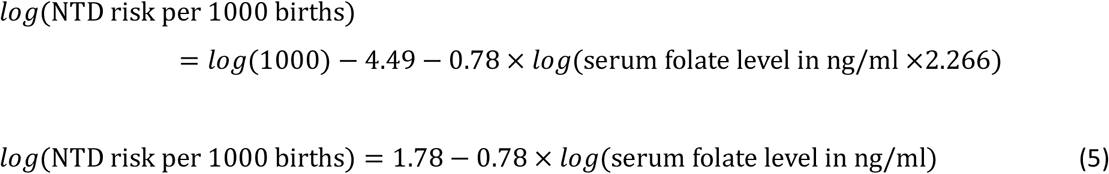
iii. The predicted decrease in risk using the results from Crider/Chen can be found by substituting an 8.8-fold increase in serum folate level using the value of *B* from Equation (5) into Equation (2):

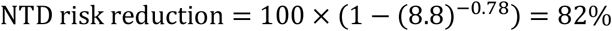

